# Supermarket fruit and vegetable placement trial: outcomes on store sales, customer purchasing, diet and household waste in a prospective matched-controlled cluster study

**DOI:** 10.1101/2025.03.17.25324102

**Authors:** Christina Vogel, Sarah Crozier, Preeti Dhuria, Joanne Lord, Graham Moon, Wendy Lawrence, Janet Cade, Kylie Ball, Cyrus Cooper, Janis Baird

**Author notes:** **Correspondence to:** Christina Vogel, Centre for Food Policy, City St George’s, University of London, Myddelton Street, London, UK. **Data sharing** Data described in this manuscript that has been collected by the research team during this study and, that which can be anonymised, will be made available upon reasonable request to the data manager (Vanessa Cox,) pending their approval. Purchasing and sales data are confidential and will not be made available because of the conditions of the agreement with the commercial collaborator. **Contributor statement** CV and JB conceived the study. CV and SC wrote the first draft of the manuscript. CV, JB, SC, KB, GM, JL, WL, JC and CC designed the study and the research methodologies. CV, JB, PD, and SC developed the data collection tools. PD, CV and SC collected and cleaned the data. SC conducted the analyses with input from CV and JB. All authors helped to draft the final manuscript. **Trial registration** NCT03573973; Pre-results.

## Abstract

**Background:** Previous product placement trials have been underpowered and limited in outcomes. This study assessed effects of positioning an expanded fruit and vegetable section near entrances on store-level sales, household-level purchasing and waste, and dietary behaviours.

**Methods:** This prospective matched controlled cluster trial (NIHR 17/44/46) involved 36 stores (18 intervention and 18 control) of a discount supermarket chain in England. The intervention was implemented for six months. Control stores were matched on store sales, customer profiles and neighbourhood deprivation. Women customers aged 18 to 60 years, with loyalty cards, were assigned to the intervention (n=280) or control group (n=300).

**Results:** Interrupted time series analyses showed increases in store-level sales of fruit and vegetables were greater in intervention stores than predicted at intervention implementation (0.32SDs (95%CI 0.11, 0.53), p=0.002) and 3-months (0.23SDs (95%CI −0.05, 0.52), p=0.10) and 6-months follow-up (0.18SDs (−0.16, 0.52), p=0.29), equivalent to ∼2525 (95% CI 775, 4115), 1940 (95% CI 380, 3950) and 1450 (95% CI −945, 3950) extra portions per store, per week respectively. Effect sizes were somewhat stronger in stores where the produce section moved forward further. Proportion of households purchasing fruit and vegetables were somewhat protected among intervention compared to control participants. Changes in dietary quality were small but generally in the direction for health benefit. Change in frequency of household vegetable waste was negligible at 3-month follow-up but increased at 6-months.

**Interpretation:** Positioning produce sections near supermarket entrances can improve the nutrition profile of store sales and may improve household purchasing and dietary quality.

**Trial registration:** NCT03573973.

**Research in Context:** *Evidence before this study:*

- Obesity and poor diet are major public health concerns and retailers’ marketing strategies impact food choices
- Product placement is a marketing strategy used by many retailers to promote unhealthy foods
- Governments are beginning to ban the placement of unhealthy foods at locations such as store entrances, checkouts and aisle ends in large retailers, in-store and online

*Added value of this study:*

- Research from adequately powered, robustly designed real-world studies on the prominent placement of healthy foods can inform improvements to existing regulations to maximise their impact on population diet
- Positioning an expanded fruit and vegetable section near store entrances increased fresh and vegetable sales at the population level; this intervention may protect against declines in household produce purchasing, particularly when exposed to a greater intervention dose

*Implications of all the available evidence:*

- Government regulations to curb placement marketing strategies being used by retailers to promote unhealthy foods should consider requiring the placement of a fruit and vegetables section at store entrances alongside limiting placement of unhealthy foods in locations such as checkouts, aisle-ends and store entrances to maximise their health

## 1. Introduction

Obesity and poor diet constitute two of the greatest threats to population health. They are key priorities of government policies because they contribute directly to poor productivity, mortality and health inequalities [1]. There are widening inequalities in diet, weight status and life expectancy between socioeconomically vulnerable and affluent families [2–3].

The food industry and citizens are trapped in a ‘*junk food cycle’*, where unhealthy foods are cheap to make, profitable to market, appealing to eat and affordable to buy [4]. Healthy food is twice as expensive per calorie than unhealthy foods high in fat, sugar and salt (HFSS), and <1% of placement promotions are for fruit and vegetables [5–6]. In recognition of the need for policies that help shape food environments to support everyone to achieve and sustain a healthy diet, the UK Government implemented the first component of the Food (Promotions and Placement) regulations on 1^st^ October 2022 in England [7]. This law supports the creation of healthier store layouts in all large retailers by restricting the positioning of HFSS products in prominent in-store locations including store entrances, aisle-ends and checkouts. A pre-implementation assessment of this regulation indicated widespread support for these new rules, however, some stakeholders felt the restrictions could go further in supporting everyone to make healthier food choices [8–9].

Reviews of the scientific literature assessing the effectiveness of product placement strategies indicate moderate evidence for healthier positioning strategies improving diet-related behaviours such as store sales, household purchasing and dietary patterns. Effects are stronger when healthy foods are positioned in prominent locations and unhealthy foods are concurrently positioned in less prominent locations, and there is greater consistency of effects when healthier positioning strategies are combined with increasing availability of healthy foods or reducing availability of unhealthy products [10–11]. Few studies in this field of research are adequately powered due to methodological difficulties implementing robust study designs in real-world retail settings [10, 12]. There is also limited evidence of the differential effects by socioeconomic status, yet this information is particularly important for policy makers [12–13].

While many supermarkets do place fresh fruit and vegetables in a position that customers encounter when first entering the store, discount and small supermarket chains do not routinely place fruit and vegetables near the store entrance. UK research shows that discount and small supermarkets have less healthy environments than other UK supermarkets, including lower availability and less prominent placement of fresh fruit and vegetables [14]. These poorer in-store environments may be contributing to dietary inequalities because families experiencing disadvantage and younger adults, known to have poorer quality diets, frequently rely on these stores for their food [15–16]. Evaluation of the effects of this placement strategy could inform future improvements to existing government regulations.

This study addresses several evidence gaps regarding the use of placement strategies to improve population diet. It aims to assess whether enhancing the positioning and availability of the fresh produce section in discount supermarkets in England improves the healthiness of store sales (population level, secondary outcome), household purchasing (household level, primary outcome), and dietary quality among women customers aged 18-60 years and their young children (individual level, secondary outcomes) after 3- and 6-months. This study is unique in its analysis of individual loyalty card data, in addition to store sales data, as well as collecting dietary data from more than one family member [17–18]. The study also assessed household fruit and vegetables waste patterns to provide a wholistic evaluation.

## 2. Methods

### 2.1 Study design and setting

The WRAPPED study was a natural experiment with a prospective matched controlled cluster design, with participants clustered within 36 study supermarkets to account for the store-based intervention. The flow diagram, Supplementary figure 4, illustrates the sampling frame for all outcome data points: store sales, customer purchasing, dietary data and household waste. The study took place between March 2018 and May 2022 and was registered with ClinicalTrials.gov (NCT03573973, pre-results). It was approved by the University of Southampton, Faculty of Medicine Ethics Committee (ID 20986.A9) and conducted in accordance with the Declaration of Helsinki and Data Protection regulations.

The study setting was discount supermarket stores in England. The collaborating supermarket has over 1000 stores nationwide and holds ∼2% of the UK grocery market [19].

This study sampled 36 stores, 18 intervention and 18 control stores [20]. Intervention stores were selected as those for which structural changes to their in-store environment was planned during the study period by the company. Control stores were matched to an intervention store on: i) sales profile, ii) customer profile, and iii) neighbourhood deprivation (IMD) [12, 21]. Control stores were geographically distant from intervention stores to reduce contamination effects.

### 2.2 Intervention and control conditions

The intervention was implemented continuously for 6 months with two components executed simultaneously: i) more prominent positioning of fresh fruit and vegetables near store entrances, and ii) expanding the numbers of fresh fruit and vegetables available. The positioning or availability of frozen and canned fruit and vegetables were not altered.

The control condition was the previous layout of stores with a limited range of fresh fruit and vegetables, placed at the back of the store and other products in their usual locations.

### 2.3. Participant eligibility and recruitment

Women were the target population because they act as household food gatekeepers and influence their family’s food choices [22]. Eligible participants were women aged 18–60 years, holding a study supermarket loyalty card and had shopped in a study store 12 weeks before recruitment. Women under 18 or over 60 years of age, without a loyalty card or shopped online only were not eligible.

Recruitment occurred in five waves between May 2018 and October 2021 with each pair of stores recruited over the same pre-implementation period. Eligible women, identified from loyalty card data, were sent a letter inviting them to participate. Participation involved completing four telephone interviews with a researcher at baseline and 1-, 3- and 6-months after intervention implementation, and consenting to share their loyalty card data. The letter did not contain details about the intervention and was sent by the supermarket to comply with data protection laws. Interested women contacted the research team directly via Freephone, text or email, were screened for eligibility and provided informed consent. To boost participant numbers, in-store recruitment was employed. The research team approached women customers while shopping in study stores and offered an information sheet. Interested women were subsequently phoned and consented. This method proved effective at enhancing sample diversity and was used for all 36 stores. Participants were offered up to 3x £10 Love2Shop or Amazon vouchers for their time.

Due to recruitment challenges during COVID-19, a sixth wave of recruitment was undertaken between September 2022 and February 2023 to boost purchasing data numbers (primary outcome) according to post-hoc sample size calculations (Supplementary Box 1). A letter was sent by the collaborating supermarket chain. Participants were offered a £15 Love2Shop or Amazon voucher for completing one telephone interview and consenting to share their loyalty card data.

All participants completed a demographic questionnaire about their age, ethnicity, marital status, highest educational qualification, employment status, number of children, weekly money spent on groceries and whether the study supermarket chain provided most of their household’s food. Participants provided home postcodes to identify neighbourhood deprivation status (IMD).

### 2.4 Outcome measures

Data collected included 9-months continuous *store sale* transaction data and *participant loyalty card purchasing* data (primary outcome) from the collaborating supermarket covering three time periods: i) baseline (3-months prior to intervention implementation); ii) short-term intervention effects (0-3 months post intervention commencement) to represent habit formation [23]; and iii) longer-term intervention effects (3-6 months post intervention commencement) to assess sustained changes. To understand intervention effects on household members’ diets, interview-administered telephone questionnaires obtained information about participants’ and their child’s diet aged 2-6 years (where applicable) at three time points: baseline, and 3-months and 6-months following intervention implementation. Frequency of household fruit and vegetable waste were also collected at these time points.

Store sales of fresh fruit and vegetables were provided as numbers of items for each product sold in each study week. Participant purchasing data covering the same categories were provided as the number of items for each product purchased at each store visit during the study period. Purchasing data were aggregated to present data as items per household per week. Store closure for structural changes to intervention stores affected one to four weeks of sales and purchasing data; these data were removed from the analysis for both intervention and control groups. Store sales and individual purchasing datasets consisted of 11-14 weeks before the intervention and 24-28 weeks afterwards.

Measures of women’s and children’s dietary quality were assessed using published, validated tools [24–25]. Participants were asked how often in the previous month they (or their child) consumed each of the 20 Food Frequency Questionnaire (FFQ) foods. A dietary quality score for each woman or child was calculated by multiplying their standardised reported frequency of consumption of each FFQ food by corresponding weightings derived from published tools and summing the results. Dietary quality scores were standardised (mean=0, SD=1). Higher scores represent better dietary quality characterised by higher intakes of vegetables, fruit, water and wholegrain bread and lower intakes of white bread, processed meats, chips, crisps and sugar. Household waste data used the same frequency measure as the FFQ and were recorded separately for fruit and vegetables.

### 2.5 Statistical analysis

Descriptive variables are given as percentage (frequency) for categorical variables and median (interquartile range) for non-normally distributed continuous variables. Differences between intervention and control participants were tested using chi-squared tests for categorical variables and Mann-Whitney rank sum tests for non-normally distributed continuous variables.

Store sales data were analysed using an Interrupted Time Series [26] (full details Supplementary Box 2). Weekly sales per store were transformed to normal distributions using Fisher-Yates transformations [27] to protect commercially sensitive sales figures (SDs). The time series models were fitted separately to account for store pairing followed by random effects meta-analysis [28] to synthesize differences between pairs of stores at the time of intervention, and 3- and 6-months post-intervention. This method enabled: i) retention of the store pairing design, ii) comparisons between pairs and iii) overall statements of study effect size and precision [29]. Results were interpreted on the original items sold per week scale by calculating the equivalent change on the original scale to the change from the median on the Fisher-Yates transformed scale. The collaborating supermarket chain sells only packaged fruit/vegetables (not singly), with each item averaging 5 portions (∼400g). This information informed conversions from items to portions. Process evaluation results identified the need for additional analyses including stratifying by: i) distance fresh fruit and vegetables moved forwards in the store (>14m vs < 14m), ii) post-intervention position of fresh fruit and vegetables (first/last half of first aisle), and iii) post-intervention availability of fresh fruit and vegetables (≥73/<73 stock-keeping unit (SKU) codes).

For individual purchasing data time series analysis was not possible because data were right-hand skewed (i.e. 67% of women’s weekly purchases had no fresh fruit or vegetables). Outcome data were dichotomised to indicate whether each week resulted in any study food purchases. A difference-in-difference approach was used [30] (full details Supplementary Box 2). Women’s data were analysed according to their recruitment store to conform with intention-to-treat analysis. Process evaluation results identified the need for the same additional analyses as the store sales data, plus study supermarket as main food shop. Intervention effects according to woman’s educational level (up to 16 years of age or beyond 16 years of age) were analysed according to the study protocol.

Intervention effects on changes in dietary quality from baseline to 3- and 6-months post-intervention were assessed using linear regression models. Diet was the outcome and intervention group and diet at baseline were predictors. A second set of regression models included confounding variables (age, money spent on groceries, number of children in the household and woman’s education) determined before analyses using directed acyclic graphs (DAG) (Supplementary Figures 1 and 2) [31]. Regression models were fitted in each pair of stores separately and random effects meta-analysis [28] used to synthesize results. Intervention effects on household fruit and vegetable waste per week was assessed using the same approach (Supplementary Figure 3).

Analyses were performed in Stata 14 [32], except the time series models which were fitted in R [33]. Consistent with current statistical thinking [34] our interpretations focus on effect sizes, and their precision, rather than emphasising statistical significance relative to any boundary.

## 3. Results

### 3.1 Participant characteristics

A total of 580 women were recruited; 300 were from control stores and 280 from intervention stores (Supplementary Figure 4). Most women provided purchasing data (n=475, 248 from control stores and 227 from intervention stores which provides 85% power at 5% significance level (2-sided) (Supplementary Box 1)) and 360 women provided information about their dietary and household fruit and vegetable waste patterns (190 from control stores and 170 from intervention stores). Of these 360 women, 250 reported living with children (aged <18 years) and 127 provided data about their child aged 2-6 years. Attrition rates for diet and waste data were 13% at 3-month and 19% at 6-month follow-ups. There were slight differences in participant characteristics at baseline between intervention and control participants, with intervention women less likely to identify as being of white ethnicity and more likely to live in more deprived neighbourhoods (Table 1). More than half of the sample were aged 31-45 years, 72% identified as being white British, 40% had low educational attainment (no qualifications beyond age 16 years) and 63% were in paid employment. Almost a third reported that the study supermarket chain was where they purchased most of their groceries (31%).

**Table 1.** Baseline characteristics of all participants (n=580)

| **Characteristic** | **Total**<br>**(n = 580)** | **Control**<br>**(n = 300)** | **Intervention**<br>**(n = 280)** | **P-value** |
| --- | --- | --- | --- | --- |
| Age group, % (n) |  |  |  | 0.45 |
| 18-30 years | 16% (90) | 17% (51) | 14% (39) |  |
| 31-45 years | 51% (298) | 50% (151) | 53% (147) |  |
| 46-60 years | 33% (192) | 33% (98) | 34% (94) |  |
| White ethnicity, % (n) | 72% (417) | 80% (239) | 64% (178) | < 0.001 |
| Married/civil partnership/cohabiting, % (n) | 61% (354) | 66% (197) | 56% (157) | 0.05 |
| Low education (no qualifications beyond age 16), % (n) | 40% (224) | 40% (117) | 39% (107) | 0.36 |
| Most deprived half of neighbourhood deprivation (IMD), % (n) | 69% (400) | 61% (183) | 78% (217) | <0.001 |
| Paid employment, % (n) | 63% (361) | 66% (193) | 60% (168) | 0.16 |
| Main supermarket used is study supermarket, % (n) | 31% (177) | 29% (85) | 33% (92) | 0.25 |
| Pounds (£) spent on food per week, median (IQR) | 70 (50, 100) | 70 (50, 100) | 70 (50, 100) | 0.74 |
| Number of children in the house, median (IQR) | 1 (0, 2) | 1 (0, 2) | 1 (0, 2) | 0.30 |

### 3.2 Store sales

The synthesised results using meta-analysis are shown in Figures 1-3, illustrating results at baseline, and 3- and 6-months post intervention implementation respectively. Increases in sales of fresh fruit and vegetables were greater in the intervention stores than would be predicted by the model counterfactuals at the time of intervention (difference = 0.32 SDs (95% CI 0.11, 0.53), P=0.002), with a reduction in effect at 3-months post-intervention (difference = 0.23 SDs (95% CI −0.05, 0.52), P=0.10) and 6-months post-intervention (difference = 0.18 SDs (95% CI −0.16, 0.52), P=0.29). These changes are approximately equivalent to 2525 (95% CI 775, 4115), 1940 (95% CI 380, 3950) and 1450 (95% CI −945, 3950) extra fruit and vegetable portions per store, per week at intervention, and 3-and 6-months post intervention implementation respectively.

**Figure 1.**
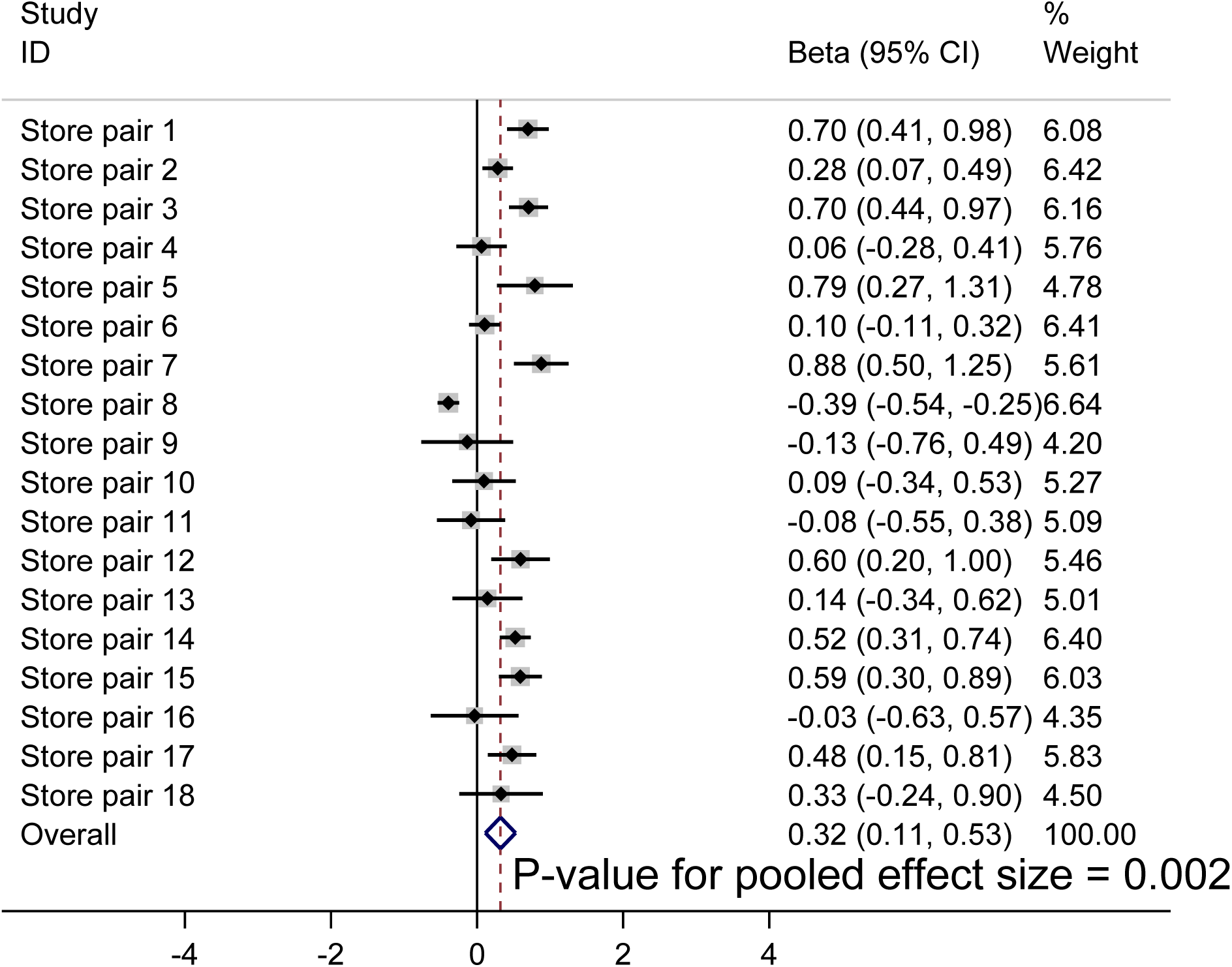
Meta-analysis of increase in store sales of fresh fruit and vegetables (SDs) in intervention stores compared to that predicted by model counterfactuals at intervention implementation.

**Figure 2.**
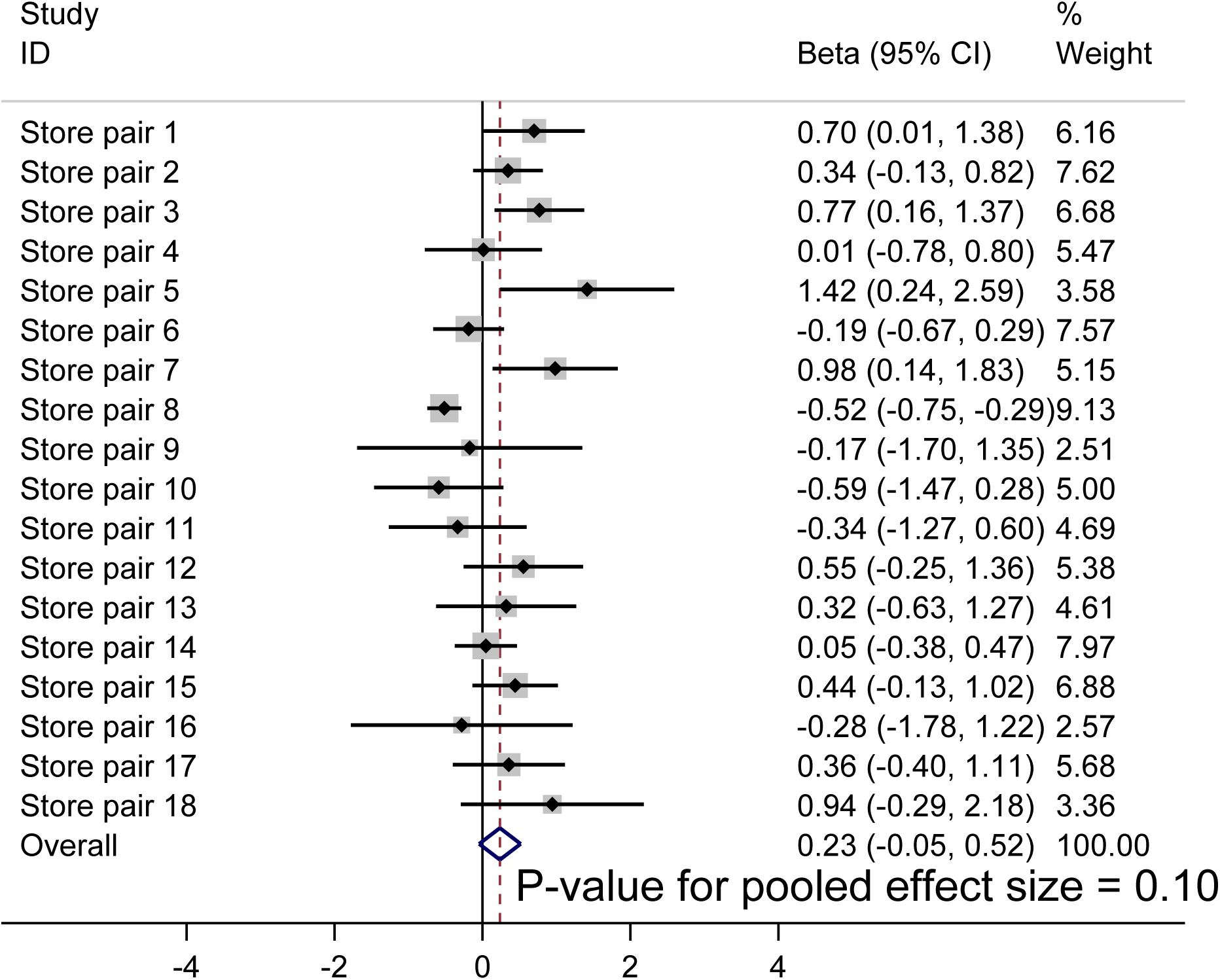
Meta-analysis of increase in store sales of fresh fruit and vegetables (SDs) in intervention stores compared to that predicted by model counterfactuals at 3-months post intervention implementation.

**Figure 3.**
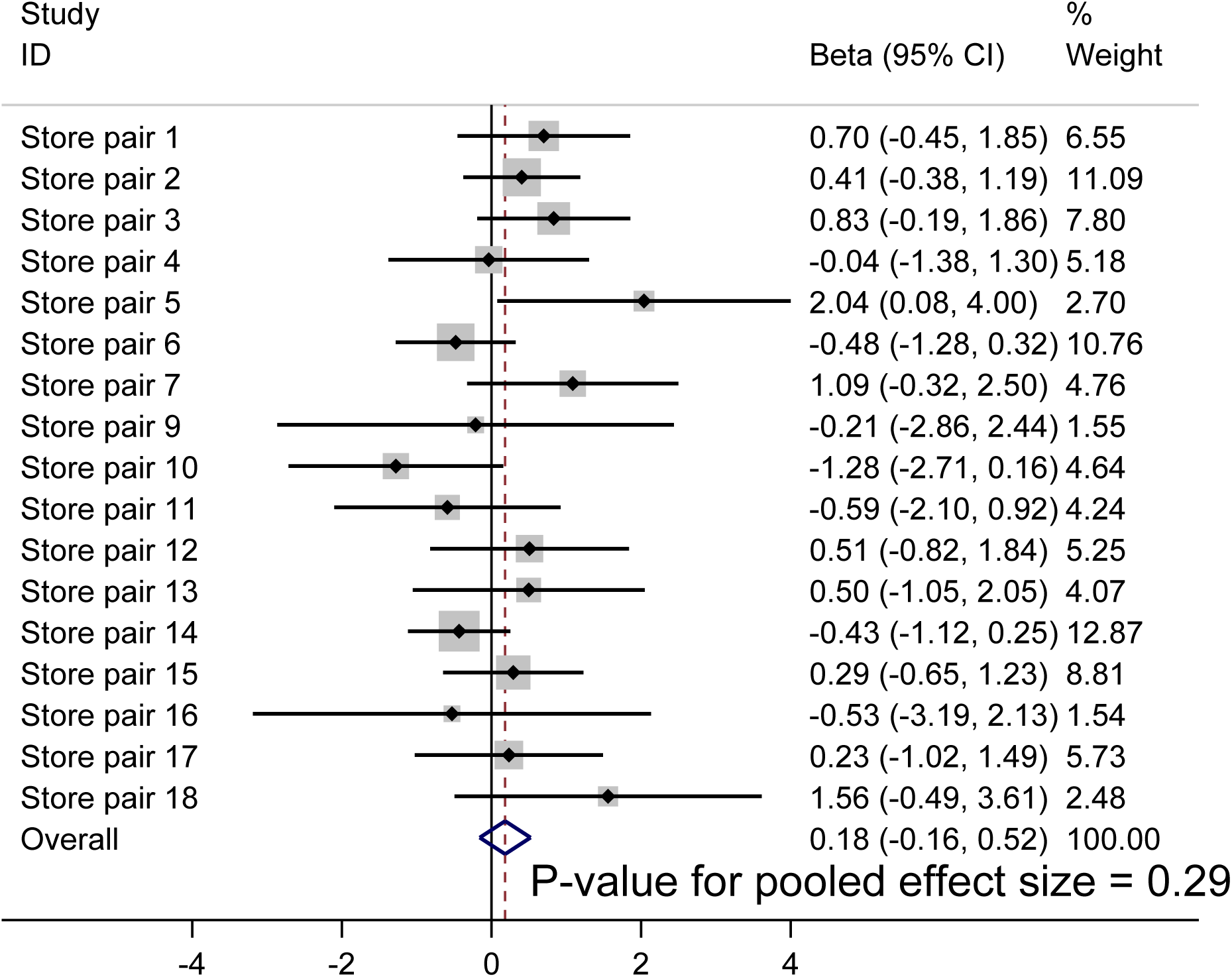
Meta-analysis of increase in store sales of fresh fruit and vegetables (SDs) in intervention stores compared to that predicted by model counterfactuals at 6-months post intervention implementation.

There were somewhat greater increases in sales of fresh fruit and vegetables in the intervention stores where the produce section moved forward more than 14 metres (Table 2; difference = 0.48 SDs (95% CI 0.30, 0.67), P<0.001 at intervention; difference = 0.40 SDs (95% CI 0.12, 0.68) at 3-months post intervention, P=0.005; difference = 0.30 SDs (95% CI − 0.13, 0.74), P=0.17 at 6-months post intervention) and lower to no increase in stores were the fruit and vegetables moved fewer than 14 metres further forward (difference = 0.15 SDs (95% CI −0.16, 0.45), P=0.35 at intervention; difference = 0.01 SDs (95% CI −0.38, 0.40) at 3-months post intervention, P=0.97; difference = −0.01 SDs (95% CI −0.57, 0.56), P=0.98 at 6-months post intervention). The changes in stores where fruit and vegetables moved forward more than 14 metres are approximately equivalent to 3645 (95% CI 2350, 5305), 3115 (95% CI 960, 5350) and 2350 (95%CI 720, 6045) greater fruit and vegetable portions per store, per week at intervention and 3- and 6-months respectively. More mixed findings were observed for intervention dose identified as being in the first, compared to the last half of the first aisle (Table 2). No notable differences in effect size were observed between stores that sold ≥73 fresh fruit and vegetables SKUs post-intervention or those which sold <73 SKUs (Supplementary Table 1).

**Table 2.** Increase in store sales of fresh fruit and vegetables (SDs) in intervention stores compared to that predicted by model counterfactuals at intervention, and 3- and 6-months follow-up post-intervention by dose (positioning)

| **Store location** | **Overall effect size SD (95% CI)** | **P-value** | **Number of stores** |
| --- | --- | --- | --- |
| **All stores** |  |  |  |
| At intervention | 0.32 (0.11, 0.53) | 0.002 | 36 |
| 12 weeks post-intervention | 0.23 (-0.05, 0.52) | 0.10 | 36 |
| 24 weeks post-intervention | 0.18 (-0.16, 0.52) | 0.29 | 36 |
| **Fruit and vegetables moved $\geq 14\text{m}$ forwards** |  |  |  |
| At intervention | 0.48 (0.30, 0.67) | < 0.001 | 18 |
| 12 weeks post-intervention | 0.40 (0.12, 0.68) | 0.005 | 18 |
| 24 weeks post-intervention | 0.30 (-0.13, 0.74) | 0.17 | 18 |
| **Fruit and vegetables moved $< 14\text{m}$ forwards** |  |  |  |
| At intervention | 0.15 (-0.16, 0.45) | 0.35 | 18 |
| 12 weeks post-intervention | 0.01 (-0.38, 0.40) | 0.97 | 18 |
| 24 weeks post-intervention | -0.01 (-0.57, 0.56) | 0.98 | 18 |
| **Fruit and vegetables to first half of first aisle** |  |  |  |
| At intervention | 0.41 (0.25, 0.57) | < 0.001 | 26 |
| 12 weeks post-intervention | 0.27 (0.02, 0.53) | 0.04 | 26 |
| 24 weeks post-intervention | 0.12 (-0.27, 0.52) | 0.53 | 26 |
| **Fruit and vegetables to last half of first aisle** |  |  |  |
| At intervention | 0.09 (-0.37, 0.56) | 0.69 | 10 |
| 12 weeks post-intervention | 0.15 (-0.52, 0.81) | 0.67 | 10 |
| 24 weeks post-intervention | 0.61 (-0.24, 1.47) | 0.16 | 10 |

### 3.3 Individual purchasing

Of the 475 participants with purchasing data, there were 5077 store visits according to their loyalty cards over the study period, 2704 from control store participants and 2373 from intervention store participants. Of the 5077 visits, 1724 (955 control women visits, 769 intervention women visits) were not at stores where women had been recruited and, while 72 of these visits were to another study store, only three visits were to stores in the opposite study arm.

Modelled proportions of women’s fruit and vegetable show an overall decline in purchasing over time (Figure 4). The proportion of control participants purchasing fruit and vegetables decreased from baseline to 3-months (−1.4% (95% CI −5.4%, 2.5%)) and from baseline to 6-months (−4.8% (95% CI −8.7%, −1.0%)); amongst intervention participants the reduction was less marked than among control participants at 3-months (−0.8% (95% CI −5.1%, 3.6%)) and at 6-months (−1.4% (95% CI −5.7%, 2.9%; Table 3). When stratifying by intervention dose using distance produce section moved, the reduction in proportion purchasing fruit and vegetables amongst intervention participants compared to control participants was less-marked (though not statistically significant) among those who shopped at stores where the produced section moved ≥14 meters compared to those where the produce section moved <14 meters (Table 3). Among participants whose main supermarket was the study store, those shopping at intervention stores showed somewhat less marked reductions in fruit and vegetable purchasing at 3-months and 6-months follow-up compared to those who shopped at control stores (Table 3). Results for stratification by highest educational qualification, as a marker of socioeconomic status, showed that the proportion purchasing fruit and vegetables amongst intervention participants increased somewhat from baseline among those with fewer educational qualifications compared to those with more educational qualifications at 6 months (Table 3). The same contrast was not observed among control participants. There were similar differences in effect size according to stratification by number of SKUs (≥73 or <73) in intervention stores at 6-months, but not for positioning of produce section in first half or last half of the aisle in intervention stores (Supplementary Table 2).

**Figure 4.**
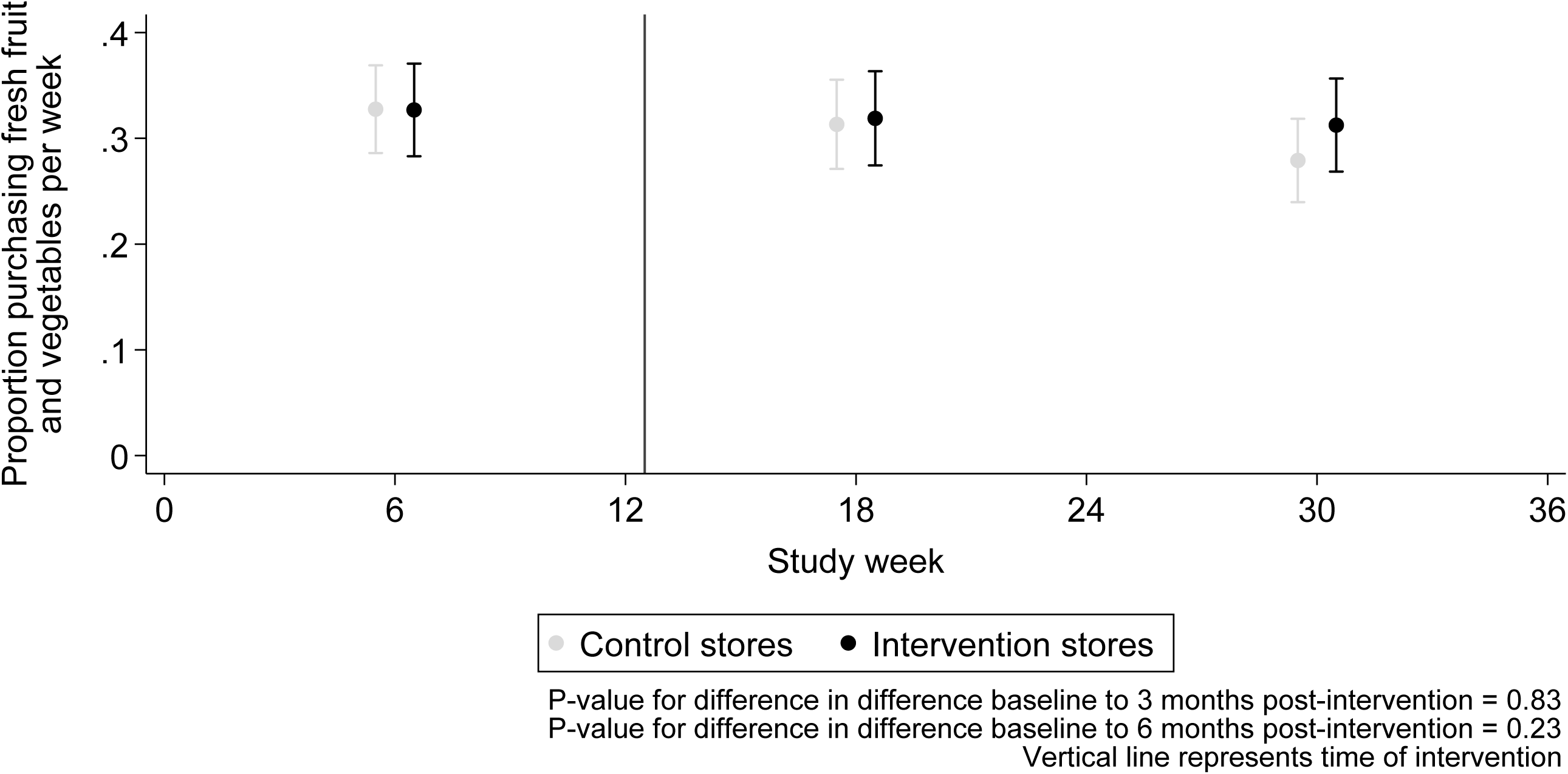
Modelled proportion of women purchasing food items in intervention and control stores.

**Table 3.** Effect of intervention on change in proportion of fresh fruit and vegetable purchasing from baseline to 3- and 6-months follow-up post-intervention.

|  | Control | Intervention | Number of stores | Number of women | Number of visits | P-value for control vs intervention | P-value for interaction |
| --- | --- | --- | --- | --- | --- | --- | --- |
|  | Change (95% CI) | Change (95% CI) |  |  |  |  |  |
| Baseline to 3 months | -1.4% (-5.4%, 2.5%) | -0.8% (-5.1%, 3.6%) | 36 | 475 | 4793 | 0.83 |  |
| Baseline to 6 months | -4.8% (-8.7%, -1.0%) | -1.4% (-5.7%, 2.9%) |  |  |  | 0.23 |  |
| Moved ≥14m forwards |  |  |  |  |  |  |  |
| Baseline to 3 months | -3.7% (-8.9%, 1.6%) | -1.6% (-7.8%, 4.7%) | 18 | 242 | 2462 | 0.61 |  |
| Baseline to 6 months | -6.1% (-11.2%, -0.9%) | -1.0% (-7.2%, 5.2%) |  |  |  | 0.20 |  |
| Moved <14m forwards |  |  |  |  |  |  |  |
| Baseline to 3 months | 1.2% (-4.8%, 7.2%) | 0.0% (-6.0%, 6.2%) | 18 | 233 | 2331 | 0.80 | 0.44 |
| Baseline to 6 months | -3.5% (-9.2%, 2.2%) | -1.8% (-7.8%, 4.2%) |  |  |  | 0.69 | 0.50 |
| Study store is main store |  |  |  |  |  |  |  |
| Baseline to 3 months | -3.5% (-10.5%, 3.4%) | 0.0% (-7.5%, 7.6%) | 33 | 138 | 1755 | 0.49 |  |
| Baseline to 6 months | -10.3% (-16.9%, -3.7%) | -1.0% (-8.5%, 6.3%) |  |  |  | 0.07 |  |
| Study store is not main store |  |  |  |  |  |  |  |
| Baseline to 3 months | -0.4% (-5.2%, 4.4%) | -1.3% (-6.6%, 4.1%) | 36 | 337 | 3038 | 0.82 | 0.49 |
| Baseline to 6 months | -2.0% (-6.8%, 2.7%) | -1.7% (-7.0%, 3.6%) |  |  |  | 0.91 | 0.18 |
| Educated up to age 16 years |  |  |  |  |  |  |  |
| Baseline to 3 months | 3.4% (-2.9%, 9.8%) | 2.8% (-4.0%, 9.8%) | 36 | 180 | 1863 | 0.94 |  |
| Baseline to 6 months | -2.7% (-8.8%, 3.3%) | 5.0% (-1.8%, 11.8%) |  |  |  | 0.09 |  |
| Educated over 16 years of age |  |  |  |  |  |  |  |
| Baseline to 3 months | -3.3% (-8.6%, 1.9%) | -2.5% (-8.2%, 3.3%) | 35 | 283 | 2812 | 0.79 | 0.32 |
| Baseline to 6 months | -5.0% (-10.3%, -0.9%) | -4.7% (-10.3%, 1.0%) |  |  |  | 0.66 | 0.19 |

### 3.4 Dietary variables

Dietary quality findings (Table 4) indicate a positive effect at 6-months post-intervention (0.25 SDs (95% CI 0.10, 0.40) among women. The effect was small, however, at 1-month follow-up (0.07 SDs (95% CI −0.09, 0.23) and in the opposite direction at 3-months follow-up (−0.15 SDs (95% CI −0.33, 0.03). Children’s dietary quality demonstrated positive effects of the intervention on dietary quality at 1- and 3-months follow-up (0.34 SDs (95% CI −0.30, 0.98) and 0.43 SDs (95% CI −0.39, 1.25) respectively), although uncertainty is indicated by wide confidence intervals. There were no effects at 6-months post-intervention (0.06 SD (95% CI −0.13, 0.24)).

**Table 4.**
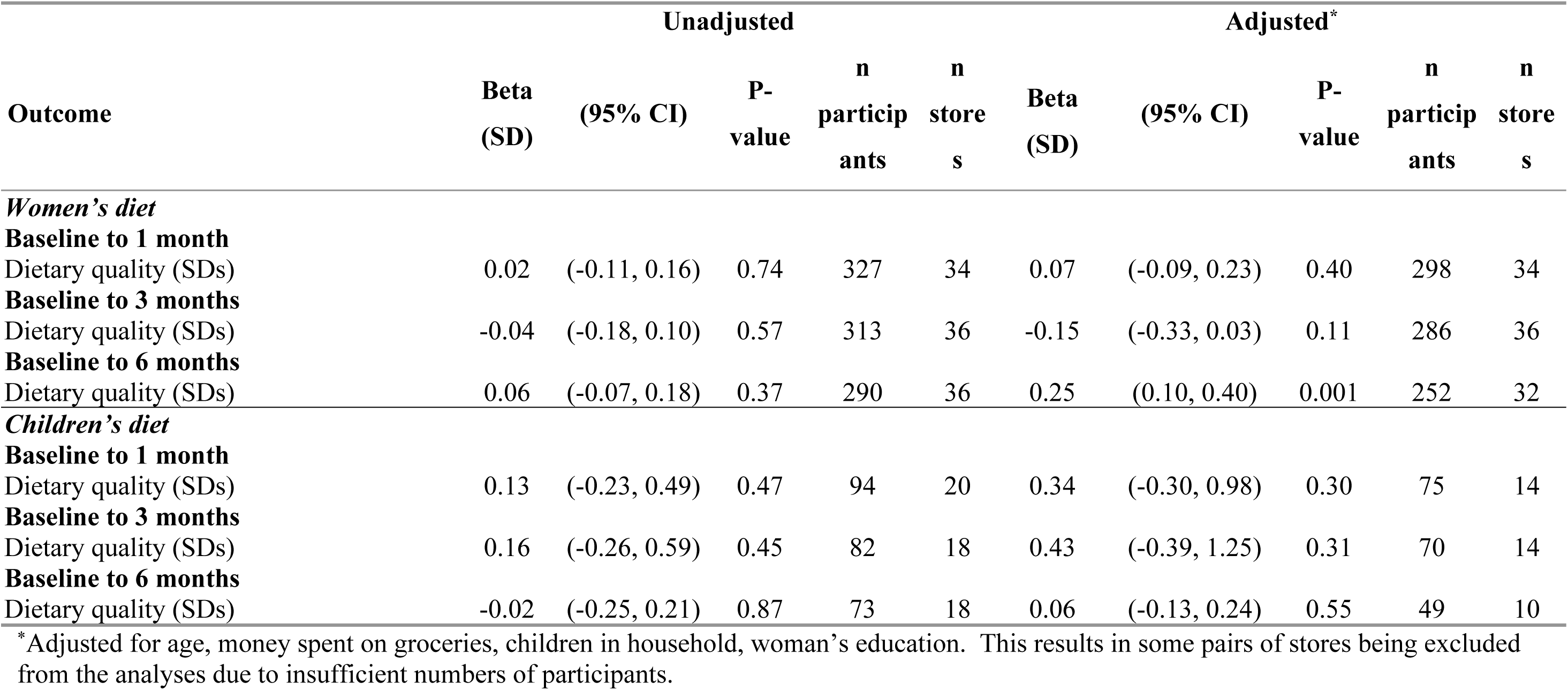
Effect of store intervention on women’s and children’s dietary quality from baseline to 1-, 3- and 6-months follow-up post-intervention.

| Outcome | Unadjusted |  |  |  |  | Adjusted* |  |  |  |  |
| --- | --- | --- | --- | --- | --- | --- | --- | --- | --- | --- |
|  | Beta (SD) | (95% CI) | P-value | n participants | n stores | Beta (SD) | (95% CI) | P-value | n participants | n stores |
| *Women's diet* |  |  |  |  |  |  |  |  |  |  |
| **Baseline to 1 month** |  |  |  |  |  |  |  |  |  |  |
| Dietary quality (SDs) | 0.02 | (-0.11, 0.16) | 0.74 | 327 | 34 | 0.07 | (-0.09, 0.23) | 0.40 | 298 | 34 |
| **Baseline to 3 months** |  |  |  |  |  |  |  |  |  |  |
| Dietary quality (SDs) | -0.04 | (-0.18, 0.10) | 0.57 | 313 | 36 | -0.15 | (-0.33, 0.03) | 0.11 | 286 | 36 |
| **Baseline to 6 months** |  |  |  |  |  |  |  |  |  |  |
| Dietary quality (SDs) | 0.06 | (-0.07, 0.18) | 0.37 | 290 | 36 | 0.25 | (0.10, 0.40) | 0.001 | 252 | 32 |
| *Children's diet* |  |  |  |  |  |  |  |  |  |  |
| **Baseline to 1 month** |  |  |  |  |  |  |  |  |  |  |
| Dietary quality (SDs) | 0.13 | (-0.23, 0.49) | 0.47 | 94 | 20 | 0.34 | (-0.30, 0.98) | 0.30 | 75 | 14 |
| **Baseline to 3 months** |  |  |  |  |  |  |  |  |  |  |
| Dietary quality (SDs) | 0.16 | (-0.26, 0.59) | 0.45 | 82 | 18 | 0.43 | (-0.39, 1.25) | 0.31 | 70 | 14 |
| **Baseline to 6 months** |  |  |  |  |  |  |  |  |  |  |
| Dietary quality (SDs) | -0.02 | (-0.25, 0.21) | 0.87 | 73 | 18 | 0.06 | (-0.13, 0.24) | 0.55 | 49 | 10 |

\*Adjusted for age, money spent on groceries, children in household, woman's education. This results in some pairs of stores being excluded from the analyses due to insufficient numbers of participants.

**Table 5.**
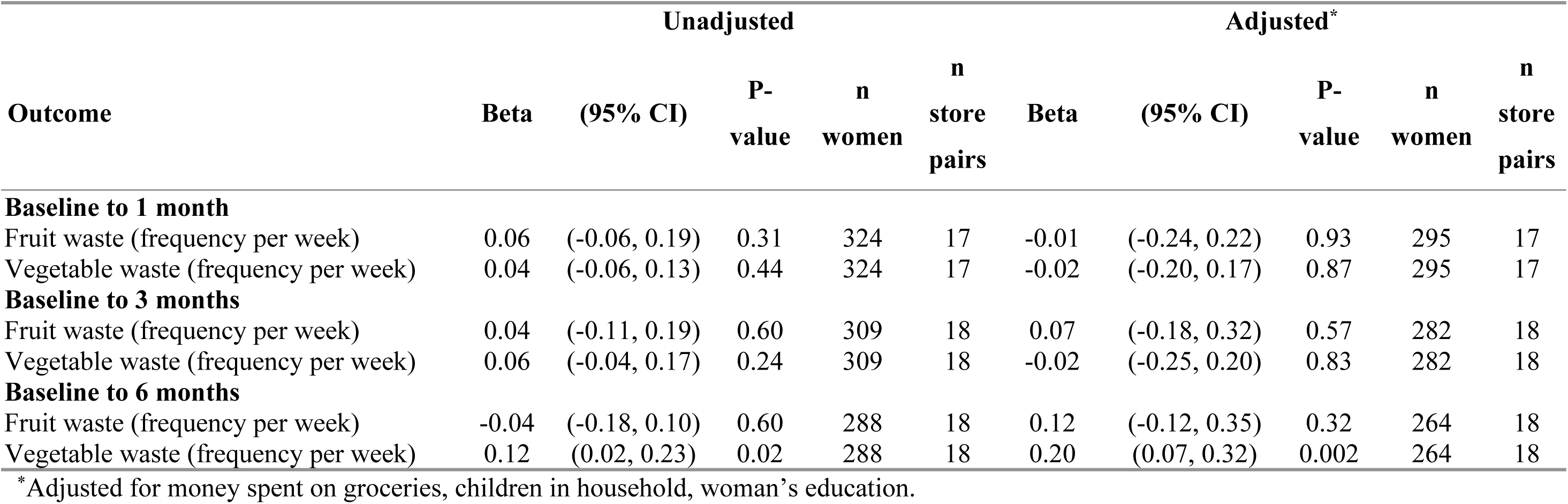
Effect of store intervention on change in frequency of waste from baseline to 1-, 3- and 6-months follow-up post-intervention.

### 3.5 Household waste

The effect of the intervention on reported household waste revealed little change in frequency of fresh fruit being thrown away at 1- (−0.01 times per week (95% CI −0.24, 0.22)) and 3- months follow-up (0.07 times per week (95% CI −0.18, 0.32)). Similarly, there was little effect on change in frequency of fresh vegetables being thrown away at 1- (−0.02 times per week (95% CI −0.20, 0.17) and 3-months follow-up ((−0.02 times per week (95% CI −0.25, 0.20)). At 6-months post-intervention, a greater frequency of vegetable waste was more evident among women from intervention than control stores (0.20 times per week (95% CI 0.07, 0.32) and fruit waste was also somewhat greater among women from intervention than control stores (0.12 times per week (95% CI −0.12, 0.35).

## 4. Discussion

This trial in discount supermarkets to enhance the placement (positioning and availability) of fresh fruit and vegetables resulted in increased fresh fruit and vegetable sales at a population (store) level, particularly when the intervention began and when produce moved further forward. Among stores with the higher intervention dose approximately 3645, 3115 and 2350 additional portions of fruit and vegetables were sold in each intervention store, per week at the time of intervention implementation, and 3- and 6-months later respectively. At the household level, the intervention indicated a small beneficial effect on fresh fruit and vegetable purchasing patterns, particularly after 6 months of intervention implementation, with a less marked reduction in fruit and vegetable purchasing among intervention compared to control families at a time when fruit and vegetable purchasing was declining across the UK population. Families using the study store for most of their groceries and those experiencing low socioeconomic position obtained slightly greater benefit from the intervention with a higher proportion of intervention families purchasing fresh fruit and vegetables compared to control families and these differences increasing over time. Women’s dietary quality improved after 6 months of exposure to the intervention, compared to women not exposed to the intervention, although beneficial effects of the intervention were not observed for diet at earlier time points. Children’s dietary quality improved somewhat after the intervention, but the effect size declined at 6-months. Self-reported household waste of fruit and vegetables was not different between intervention and control families immediately and shortly after intervention exposure, however, intervention families reported higher vegetable waste after 6 months compared to baseline.

### 4.1 Strengths and limitations

To our knowledge, this is the first fresh fruit and vegetable placement trial in supermarkets to provide 85% power (Supplementary Box 1) to detect an effect of household purchasing patterns using loyalty card data and include multiple datasets to provide a thorough overview of intervention effects. Other advantages over existing research [12, 18] includes using: matched comparison group; robust statistical methods; and assessment of differential interventions effects by socioeconomic status and intervention dose.

This study limitations not being able to randomise stores potentially biasing results through unmeasured confounding effects. Nonetheless, the findings provide knowledge of intervention effectiveness in complex social contexts which is useful for policymakers. The primary outcome was adequately powered but interaction analyses were not and are harder to detect statistically [35]. Individual dietary and household waste results should be viewed cautiously due to difficulties recruiting over COVID-19 resulting in smaller than predicted samples. Store waste figures were not collected, and data from other supermarkets participants used were not available. Store selection and intervention implementation was not within the researchers’ control and some deviations from the protocol were identified. Under- or over-estimation of intervention effects may be possible, however, stratified analyses attempted to address variations.

### 4.3 Comparison with previous literature

Previous intervention studies which repositioned fresh fruit and vegetables to store entrances or other prominent locations in food stores found similar effects to our study [36–38]. The intervention dose (i.e. distance produce section moved and study store being main source of groceries) moderately boosted sales of fruit and vegetables at a population level and protected declining fruit and vegetable purchasing at the household level. Previous studies of lower intensity fruit and vegetable positioning interventions, like co-locating produce alongside unhealthy foods or having smaller displays, have shown no intervention effect or only very small intervention effects [36, 38, 39].

Our study highlights the worrying decline in fresh fruit and vegetable sales from retail outlets in the UK. From 2018-2022 British families experienced disruptions to produce availability and price rises caused by COVID-19, Brexit, climatic events, and war in Europe. Subsequently, UK household purchasing of fruit dropped by 7.2% and vegetable purchasing dropped by 5.3% [40]. Households are purchasing, on average, fewer than four portions of fruit and vegetables per day for the entire family. With approximately 2.4 residents in UK households it is not surprising only 33% of adults meet the recommended 5-a-day [41]. Our study shows a slight reduction in intervention effect size on store level sales 6-months post intervention implementation which mirrors population trends. Even small increases in fruit and vegetable consumption (0.3-1.0 portion/day) can reduce an individual’s risk of coronary heart disease by 4% and stroke by 5% [42–43]. Our study results illustrate potentially meaningful effect size at the population level despite contextual disruptions.

At the household level the intervention provided somewhat protective effects against these national trends, with differences in fruit and vegetable purchasing between intervention and control families increasing slightly over time. These results suggest intervention effects may be strongest at the household and individual levels after 6-months of continued intervention exposure. This effect was greater among families relying on the study store for most of their groceries and those experiencing lower socioeconomic position. Evidence from the US shows similar findings for families receiving governmental financial support to buy nutritious foods for their young children, with higher use after five months of continued exposure to produce sections near their store’s entrance [37]. The broader literature on changing health-related practices indicates that new habits take more than 12 weeks to form and strengthen with time [44], particularly if contexts or environments support those new practices [45–46].

Observational evidence shows UK discount and small supermarkets who attract families with lower incomes do not provide this contextual support, frequently <u>not</u> positioning fruit and vegetable sections near store entrances [6, 14]. Positioning produce sections in this prominent in-store location across all supermarkets could provide small protective benefits for those most at risk of low fruit and vegetable intake.

### 4.4 Implications for policy

Our study provides robust and contextually relevant considerations for food policy. The store level findings offer evidence to support expansion of the UK Food (Promotions and Placement) regulations in England requiring the positioning of fresh produce sections near store entrances in all large food stores (>2000 square feet) to boost fruit and vegetable sales and improve population diet. The international literature indicates concurrently positioning healthy foods in prominent locations and repositioning unhealthy foods to less prominent locations is more beneficial for diet-related outcomes than co-locating unhealthy and healthy items in prominent locations or implementing only one positioning strategy [10, 47]. While the UK Ofcom 2004/2005 nutrient profile model [48] used to define unhealthy foods for the Food (Promotions and Placement) regulations has prompted reformulation of savoury snacks and pizzas sold in retail outlets [49], it does not adhere to current UK dietary guidelines and still allows prominent positioning of less healthy, high-sugar foods. Policy refinement to also require the positioning of healthy foods, like fruit and vegetables, at prominent in-store locations would help maximise the regulation’s population health impact. By illustration, findings from the pilot phase of our study, which concurrently increased the placement (availability and positioning) of produce and removed unhealthy items from checkouts and aisle ends opposite (replaced with non-food items), showed an increase of approximately 9820 additional fruit and vegetable portions per store per week after 6-months [21]. Comparatively, the current full-scale study, which only improved the placement of produce, revealed an increase of approximately 1450 additional portions per store per week after 6-months. These findings demonstrate the added benefit of combining placement strategies for both healthy and unhealthy products.

## 5. Conclusion

This study provides a comprehensive assessment of a fruit and vegetable placement intervention where positioning and availability were enhanced in discount supermarkets. Findings show increased fresh fruit and vegetable sales at the population level, although effects reduced over time. Small protective effects were indicated for household fruit and vegetable purchasing and individual dietary quality because the intervention buffered against widescale reductions in fruit and vegetable purchasing caused by COVID-19 and cost-of-living challenges from 2018-2022. Somewhat greater effects were observed among families relying on study stores for their groceries and families experiencing greater socioeconomic disadvantage. This study provides novel evidence for government regulations to improve retailers’ in-store food marketing practices.

## Data Availability

Data cannot be shared publicly because of commercial and personal data sensitivities which could undermine anonymity. The non-commercial data can be made available from the MRC Lifecourse Epidemiology Centre (contact via Vanessa Cox) for researchers who meet the criteria for access to confidential data.

## Acknowledgements

We are grateful to all the head-office and store staff working at the collaborating supermarket who contributed to this study. We sincerely thank the broader WRAPPED study team for their support with data collection, particularly Julie Coleman, Julia Hammond, Karen McGill and Hannah Payne, and to Patsy Coakley for her dedicated computing and data manipulation support. We thank our funders for their support and all participants who contributed their time and information to this study.

## Supplementary Appendix

**Supplementary Figure 1:**
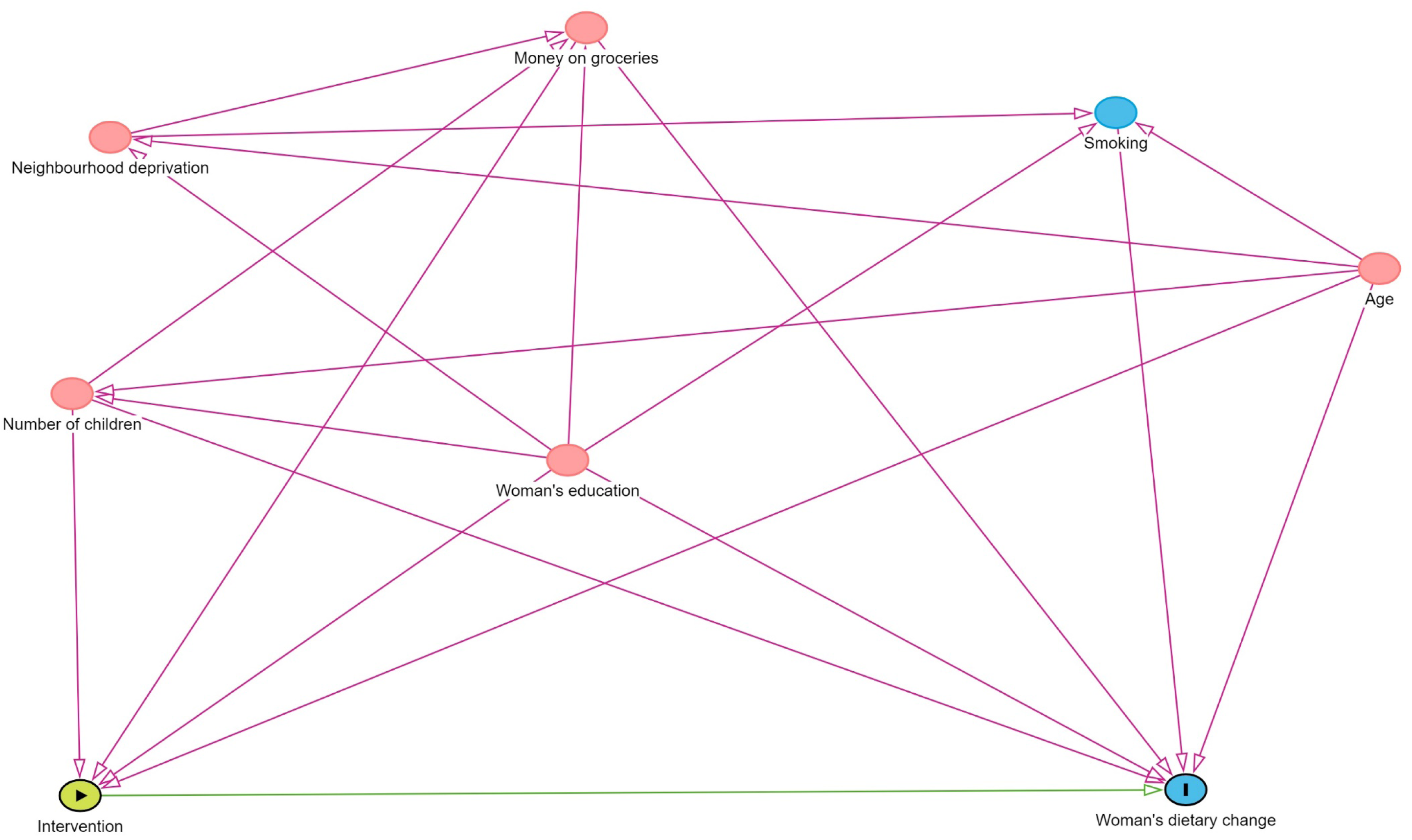
Directed Acyclic Graph for women’s dietary quality.

**Supplementary Figure 2:**
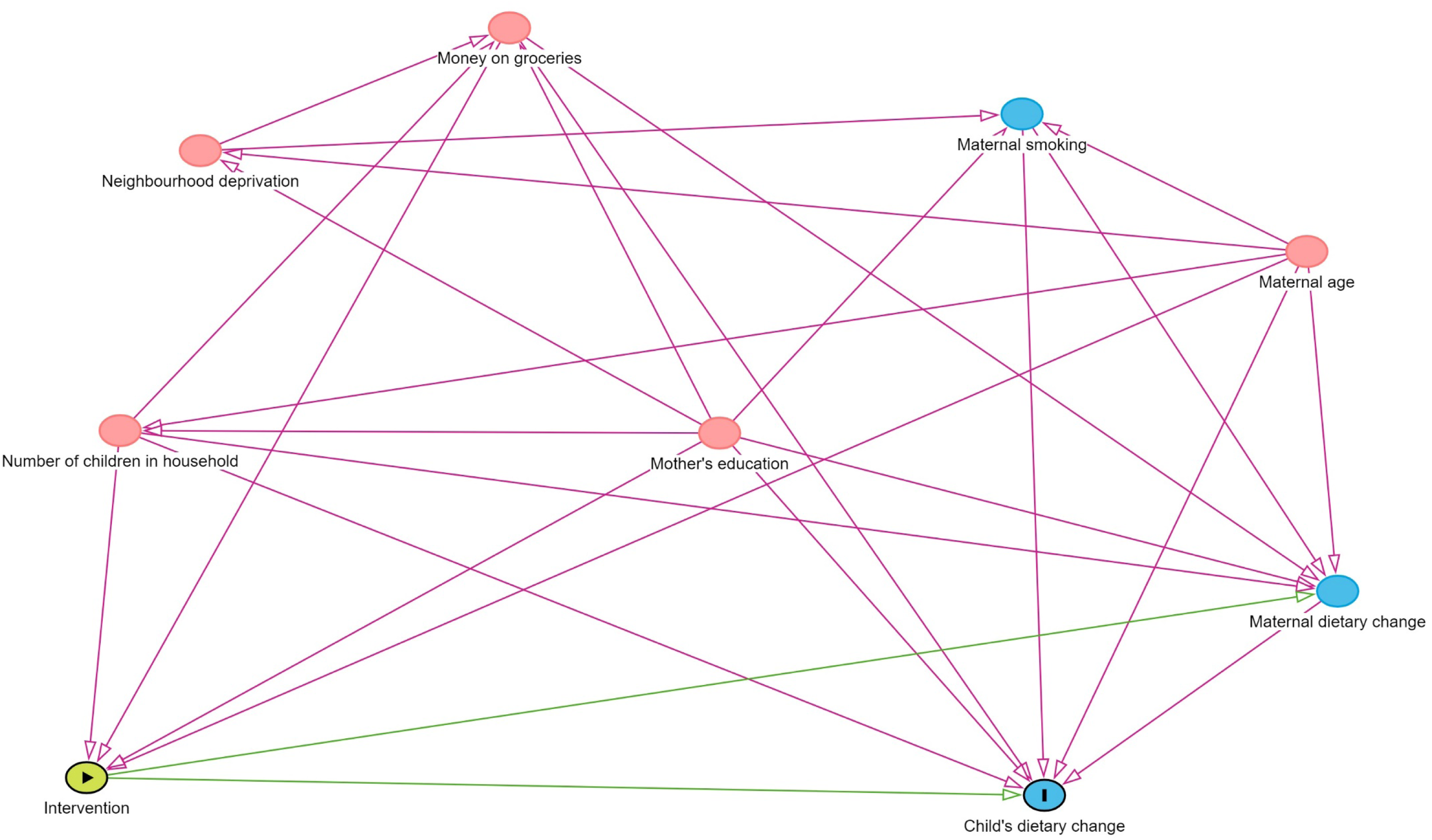
Directed Acyclic Graph for children’s dietary quality.

**Supplementary Figure 3:**
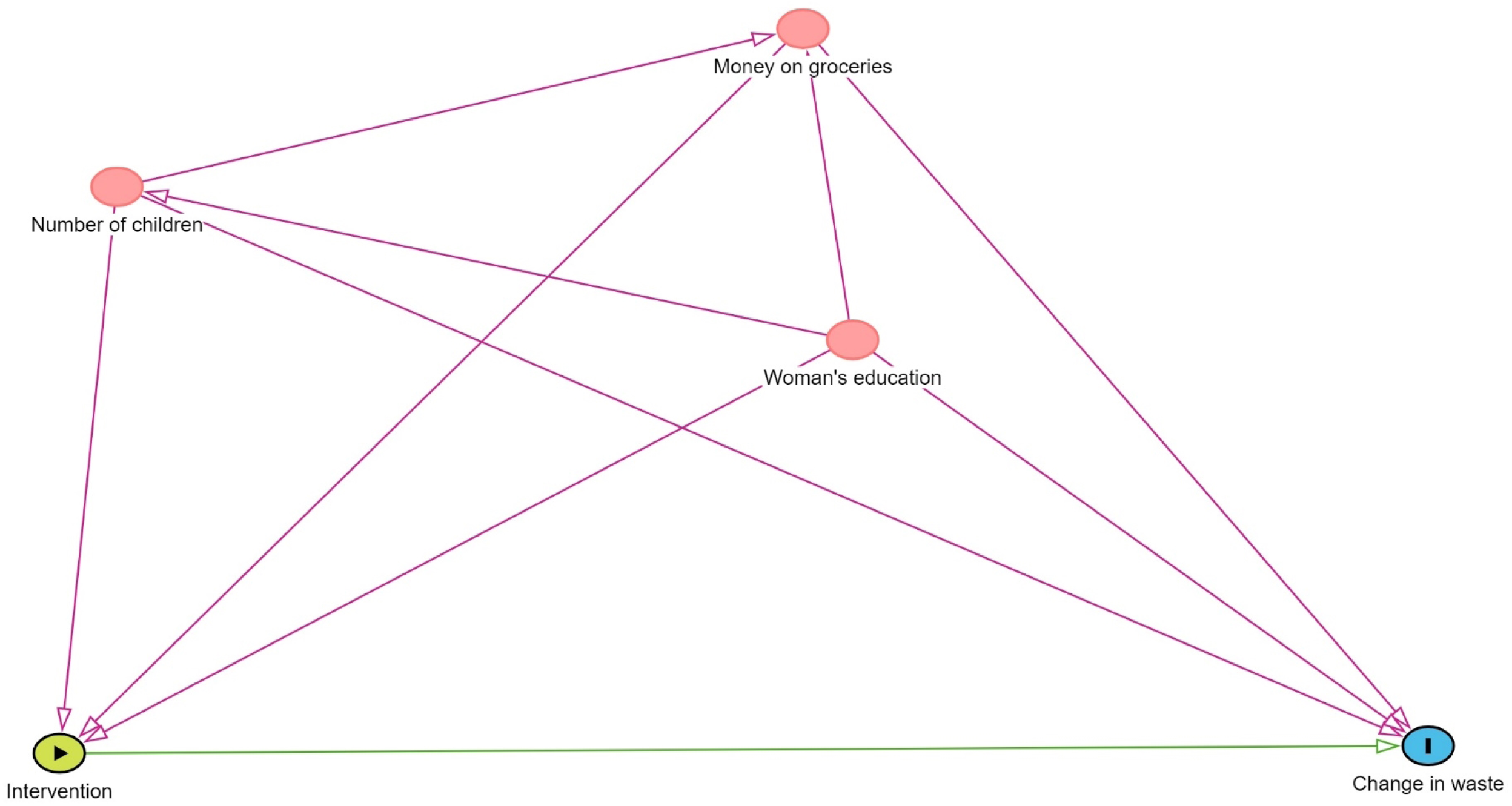
Directed Acyclic Graph for household fruit and vegetable waste.

**Supplementary Figure 4:**
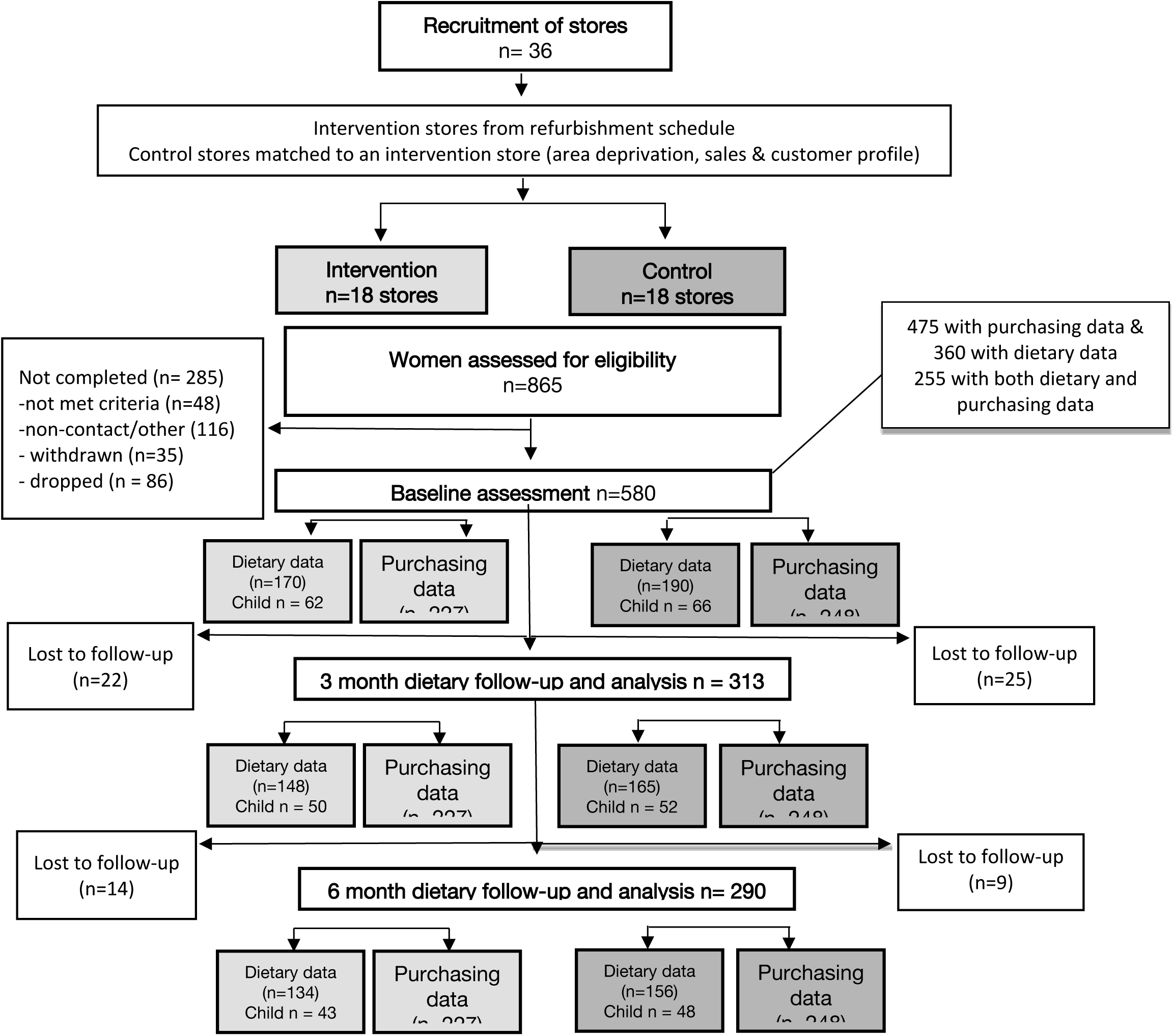
WRAPPED study flow chart and sample size.

### Supplementary Box 1

#### Sample size calculation

The study’s sample size calculations were revised post-hoc because of COVID-19 related disruptions to participant recruitment. The sixth wave of recruitment undertaken to boost participant numbers resulted in this study being powered to detect differences in the primary outcome (fresh fruit and vegetable purchasing) between women in the intervention and control groups during the 3-6 months postintervention. Average fruit and vegetable purchases per week are not normally distributed, thus the sample size calculation is based on changes in average fresh fruit and vegetable purchases per week from the baseline period to the post-intervention period, which are approximately normally distributed. It was not practical to calculate a rho from the pilot study due to the small number of clusters. We used data from our previous research on women in Hampshire who were the same age-range as the proposed participants of this study and considered the supermarkets at which the women shopped as clusters to estimate a rho of 0.1 as our intraclass correlation coefficient. We aimed to detect a difference of 0.3 items (1.5 portions) per week. Assuming a standard deviation of 0.7 items (3.5 portions) per week seen in the pilot data, 18 stores in each arm and 13 women per store (totaling 468 women) provides 85% power at a 5% significance level (2-sided).

### Supplementary Box 2

#### Interrupted Time Series analysis – store sales data

Time series models were fitted with terms for study week (linear term, weeks from baseline), intervention, level (an indicator of the post-intervention period), trend (study week in the post-intervention period), and interactions between intervention and study week, intervention and level and intervention and trend. Additional terms were included in the models for Christmas weeks, the week at the end of lockdown in June 2020 and the week before the intervention to improve model fit. By including the variable ‘level’, the model tested for a step change at the time of the intervention. The time series models were fitted separately in each pair of stores to account for store pairing. The P-value for the interaction between intervention and level indicates the significance of the impact of the intervention on store sales at the time of the intervention. Effect sizes were calculated at the 3- and 6-months post-intervention time points. A counterfactual line is included on the Interrupted Time Series graphs, indicating the trends in sales that would have been expected had the intervention not occurred. Confidence intervals at 3- and 6-months post-intervention were calculated using the delta method [32].

#### Difference in difference analysis – household purchasing data

A difference-in-difference approach was used [35] where each logistic regression model included fixed effects for intervention group, time period and the interaction between intervention group and time period. Time period was coded as two dummy variables indicating the 0-3- and 3-6-month periods post-intervention. The interaction terms tested the difference in purchasing during the intervention compared to the pre-intervention period between intervention and control stores. Random effects were included for women, to account for the multilevel structure of the data, with weeks clustered within women.

**Supplementary Table 1:** Increase in store sales of fresh fruit and vegetables (SDs) in intervention stores compared to that predicted by model counterfactuals at intervention, and 3- and 6-months follow-up post-intervention by dose (availability)

| Store location | Overall effect size 95% CI | P-value | Number of stores |
| --- | --- | --- | --- |
| All stores |  |  |  |
| At intervention | 0.32 (0.10, 0.53) | 0.004 | 36 |
| 12 weeks post-intervention | 0.23 (-0.05, 0.52) | 0.11 | 36 |
| 24 weeks post-intervention | 0.18 (-0.16, 0.52) | 0.29 | 36 |
| ≥ 73 SKU |  |  |  |
| At intervention | 0.38 (0.19, 0.57) | < 0.001 | 16 |
| 12 weeks post-intervention | 0.23 (-0.01, 0.48) | 0.06 | 16 |
| 24 weeks post-intervention | 0.02 (-0.39, 0.42) | 0.94 | 16 |
| < 73 SKU |  |  |  |
| At intervention | 0.30 (-0.01, 0.62) | 0.06 | 20 |
| 12 weeks post-intervention | 0.24 (-0.20, 0.68) | 0.28 | 20 |
| 24 weeks post-intervention | 0.31 (-0.24, 0.86) | 0.27 | 20 |

**Supplementary Table 2:** Effect of intervention on change in proportion of fresh fruit and vegetable purchasing from baseline to 3- and 6-months follow-up post-intervention stratified by dose (position and availability)

|  | Control | Intervention | Number of stores | Number of women | Number of visits | P-value for control vs intervention | P-value for interaction |
| --- | --- | --- | --- | --- | --- | --- | --- |
|  | Change (95% CI) | Change (95% CI) |  |  |  |  |  |
| **Moved first half of first aisle** |  |  |  |  |  |  |  |
| Baseline to 3 months | -1.0% (-5.5%, 3.6%) | -3.4% (-9.2%, 2.3%) | 26 | 313 | 3295 | 0.51 |  |
| Baseline to 6 months | -4.4% (-8.8%, 0.0%) | -1.0% (-6.7%, 4.7%) |  |  |  | 0.33 |  |
| **Moved last half of first aisle** |  |  |  |  |  |  |  |
| Baseline to 3 months | -3.2% (-11.2%, 4.9%) | 2.9% (-3.7%, 9.5%) | 10 | 162 | 1498 | 0.25 | 0.20 |
| Baseline to 6 months | -6.1% (-14.1%, 1.8%) | -2.2% (-8.5%, 4.2%) |  |  |  | 0.46 | 0.96 |
| **≥ 73 SKU** |  |  |  |  |  |  |  |
| Baseline to 3 months | 0.8% (-5.1%, 6.8%) | -1.0% (-7.7%, 5.7%) | 16 | 216 | 2051 | 0.69 |  |
| Baseline to 6 months | -3.0% (-8.8%, 2.8%) | 3.1% (-3.4%, 9.7%) |  |  |  | 0.17 |  |
| **< 73 SKU** |  |  |  |  |  |  |  |
| Baseline to 3 months | -3.0% (-8.3%, 2.3%) | -0.5% (-6.2%, 5.2%) | 20 | 259 | 2742 | 0.54 | 0.50 |
| Baseline to 6 months | -6.2% (-11.2%, -1.1%) | -5.2% (-10.9%, 0.4%) |  |  |  | 0.84 | 0.36 |

## References

1. Tony Blair Institute for Global Change: Unhealthy numbers: The rising cost of Obesity in the UK. In.; 2023.

2. NHS Digital: National Child Measurement Programme, England, 2022/23 School Year. In.; 2023.

3. Office for National Statistics: Health state life expectancies by national deprivation deciles, England: 2018 to 2020. In. Edited by Rea M, Tabor D. London 2022.

4. Dimbleby H: The National Food Strategy Independent Review: The Plan. In.; 2021.

5. Food Foundation: The Broken Plate 2023. In.; 2023.

6. Obesity Health Alliance: Out of place: the extent of unhealthy foods in prime locations in supermarkets. In.; 2018.

7. The Food (Promotion and Placement) (England) Regulations 2021. In: 2021 No 1368. 2021.

8. Muir S, Dhuria P, Roe E, Lawrence W, Baird J, Vogel C: UK government’s new placement legislation is a ‘good first step’: a rapid qualitative analysis of consumer, business, enforcement and health stakeholder perspectives. BMC Med 2023, 21(1):33.

9. Muir S, Dhuria P, Vogel C: Government must proceed with landmark anti-obesity regulations in England. BMJ 2022, 378:o2358.

10. Shaw SC, Ntani G, Baird J, Vogel CA: A systematic review of the influences of food store product placement on dietary-related outcomes. Nutr Rev 2020, 78(12):1030–1045.

11. Vogel C, Piernas C: The retail food environment. In: Transforming Food Environments. edn. Edited by Evans C: Routledge; 2022: 63–78.

12. Vogel C, Dijkstra C, Huitink M, Dhuria P, Poelman MP, Mackenbach JD, Crozier S, Seidell J, Baird J, Ball K: Real-life experiments in supermarkets to encourage healthy dietary-related behaviours: opportunities, challenges and lessons learned. Int J Behav Nutr Phys Act 2023, 20(1):73.

13. Cameron AJ, Charlton E, Ngan WW, Sacks G: A Systematic Review of the Effectiveness of Supermarket-Based Interventions Involving Product, Promotion, or Place on the Healthiness of Consumer Purchases. Current Nutrition Reports 2016, 5(3):129–138.

14. Black C, Ntani G, Inskip H, Cooper C, Cummins S, Moon G, Baird J: Measuring the healthfulness of food retail stores: variations by store type and neighbourhood deprivation. Int J Behav Nutr Phys Act 2014, 11(1):69.

15. Vogel C, Ntani G, Inskip H, Barker M, Cummins S, Cooper C, Moon G, Baird J: Education and the Relationship Between Supermarket Environment and Diet. Am J Prev Med 2016, 51(2):e27–e34.

16. Pechey R, Monsivais P: Supermarket Choice, Shopping Behavior, Socioeconomic Status, and Food Purchases. Am J Prev Med 2015, 49(6):868–877.

17. Appelhans BM, French SA, Tangney CC, Powell LM, Wang Y: To what extent do food purchases reflect shoppers’ diet quality and nutrient intake? Int J Behav Nutr Phys Act 2017, 14(1):46.

18. Shaw SC, Ntani G, Baird J, Vogel CA: A systematic review of the influences of food store product placement on dietary-related outcomes. Nutr Rev 2020.

19. Worldpanel K: Great Britian Grocery Market Share. In: Kantar Worldpanel Update. London; 2019.

20. Vogel C, Crozier S, Dhuria P, Shand C, Lawrence W, Cade J, Moon G, Lord J, Ball K, Cooper C, Baird J: Protocol of a natural experiment to evaluate a supermarket intervention to improve food purchasing and dietary behaviours of women (WRAPPED study) in England: a prospective matched controlled cluster design. BMJ Open 2020, 10(2):e036758.

21. Vogel C, Crozier S, Penn-Newman D, Ball K, Moon G, Lord J, Cooper C, Baird J: Altering product placement to create a healthier layout in supermarkets: Outcomes on store sales, customer purchasing, and diet in a prospective matched controlled cluster study. PLoS Med 2021, 18(9):e1003729.

22. Food Standards Agency: The 2014 Food and You survey. In. London; 2014.

23. Gardner B, Lally P, Wardle J: Making health habitual: the psychology of ‘habit-formation’ and general practice. Br J Gen Pract 2012, 62(605):664–666.

24. Crozier SR, Inskip HM, Barker ME, Lawrence WT, Cooper C, Robinson SM: Development of a 20-item food frequency questionnaire to assess a ‘prudent’ dietary pattern among young women in Southampton. European Journal of Clinical Nutrition 2010, 64(1):99–104.

25. Jarman M, Fisk CM, Ntani G, Crozier SR, Godfrey KM, Inskip HM, Cooper C, Robinson SM, Southampton Women’s Survey Study G: Assessing diets of 3-year-old children: evaluation of an FFQ. Public Health Nutr 2014, 17(5):1069–1077.

26. Penfold RB, Zhang F: Use of interrupted time series analysis in evaluating health care quality improvements. Acad Pediatr 2013, 13(6 Suppl):S38–44.

27. Armitage P, Berry G: Statistical Methods in Medical Research, Third edn. Oxford, UK: Blackwell Science Ltd; 2002.

28. Rice K, Higgins JPT, Lumley T: A re-evaluation of fixed effect(s) meta-analysis. Journal of the Royal Statistical Society: Series A (Statistics in Society) 2018, 181(1):205–227.

29. Ejlerskov KT, Sharp SJ, Stead M, Adamson AJ, White M, Adams J: Supermarket policies on less-healthy food at checkouts: Natural experimental evaluation using interrupted time series analyses of purchases. PLoS Med 2018, 15(12):e1002712.

30. Foster GD, Karpyn A, Wojtanowski AC, Davis E, Weiss S, Brensinger C, Tierney A, Guo W, Brown J, Spross C et al: Placement and promotion strategies to increase sales of healthier products in supermarkets in low-income, ethnically diverse neighborhoods: a randomized controlled trial. Am J Clin Nutr 2014, 99(6):1359–1368.

31. Greenland S, Pearl J, Robins JM: Causal diagrams for epidemiologic research. Epidemiology 1999, 10(1):37–48.

32. StataCorp: Stata: Release 14. Statistical Software. College Station, TX: StataCorp LP; 2015.

33. R Core Team: R: A language and environment for statistical computing. Vienna, Austria: R Foundation for Statistical Computing; 2013.

34. Amrhein V, Greenland S, McShane B: Scientists rise up against statistical significance. Nature 2019, 567(7748):305–307.

35. Gelman A, Hill, J., Vehtari, A., : Regression and Other Stories. In.; 2020.

36. Albert SL, Langellier BA, Sharif MZ, Chan-Golston AM, Prelip ML, Elena Garcia R, Glik DC, Belin TR, Brookmeyer R, Ortega AN: A corner store intervention to improve access to fruits and vegetables in two Latino communities. Public Health Nutr 2017, 20(12):2249–2259.

37. Thorndike AN, Bright OM, Dimond MA, Fishman R, Levy DE: Choice architecture to promote fruit and vegetable purchases by families participating in the Special Supplemental Program for Women, Infants, and Children (WIC): randomized corner store pilot study. Public Health Nutr 2017, 20(7):1297–1305.

38. Toft U, Winkler LL, Mikkelsen BE, Bloch P, Glumer C: Discounts on fruit and vegetables combined with a space management intervention increased sales in supermarkets. Eur J Clin Nutr 2017, 71(4):476–480.

39. Stuber JM, Mackenbach JD, de Bruijn G-J, Gillebaart M, Hoenink JC, Middel CNH, de Ridder DTD, van der Schouw YT, Smit EG, Velema E et al: Real-world nudging, pricing, and mobile physical activity coaching was insufficient to improve lifestyle behaviours and cardiometabolic health: the Supreme Nudge parallel cluster-randomised controlled supermarket trial. BMC Medicine 2024, 22(1):52.

40. Department for Environment Food and Rural Affairs: Family Food FYE 2022. In. London; 2023.

41. Bates B, Collins D, Cox L, Nicholson S, Page P, Roberts C, Steer T, Swan G: National Diet and Nutrition Survey, Years 1 to 9 of the Rolling Programme (2008/2009 – 2016/2017): Time trend and income analyses. In. London; 2019.

42. Dauchet L, Amouyel P, Dallongeville J: Fruit and vegetable consumption and risk of stroke: a meta-analysis of cohort studies. Neurology 2005, 65(8):1193–1197.

43. Dauchet L, Amouyel P, Hercberg S, Dallongeville J: Fruit and vegetable consumption and risk of coronary heart disease: a meta-analysis of cohort studies. J Nutr 2006, 136(10):2588–2593.

44. Gardner B, Sheals K, Wardle J, McGowan L: Putting habit into practice, and practice into habit: a process evaluation and exploration of the acceptability of a habit-based dietary behaviour change intervention. Int J Behav Nutr Phys Act 2014, 11:135.

45. Baranowski T, Missaghian M, Broadfoot A, Watson K, Cullen K, Nicklas T, Fisher J, Baranowski J, O’Donnell S: Fruit and Vegetable Shopping Practices and Social Support Scales: A Validation. Journal of Nutrition Education and Behavior 2006, 38(6):340–351.

46. Shove E, Pantzar M, Watson MF: The dynamics of social practice: Everyday life and how it changes: SAGE Publications Ltd,; 2012.

47. Vogel C, Piernas, C.: The retail food environment In: Transforming Food Environments edn. Edited by Evans C. London, UK: CRC Press; 2022.

48. Rayner M: Nutrient profiling for regulatory purposes. Proc Nutr Soc 2017, 76(3):230–236.

49. Brown R: One year on: Have HFSS rules made any difference? In: The Grocer. 2023.

